# Emergency and Sequalae Management of Traumatic Dental Injuries: A Quality Assessment of Clinical Practice Guidelines

**DOI:** 10.1101/2023.02.16.23286064

**Authors:** Carlos Zaror, Andrea Seiffert, Naira Figueiredo Deana, Gerardo Espinoza-Espinoza, Claudia Ata-la-Acevedo, Rodrigo Diaz, Alonso Carrasco-Labra

## Abstract

The prevalence and consequences of traumatic dental injuries (TDI) make them a public health problem. Trustworthy TDI Clinical Practice Guidelines (CPGs) can assist clinicians in making a proper diagnosis, and guide them to the most appropriate therapy for every case. The aim of this study was to identify and evaluate the quality of CPGs for the diagnosis, emergency management and follow-up of TDIs. A systematic search was carried out in MEDLINE, EMBASE, Epistemonikos, Trip database, CPG’ websites, and dental societies to identify documents providing recommendations for the emergency and sequelae management of TDIs. Reviewers assessed the included guidelines independently and in duplicate, using the AGREE II instrument. T-student or ANOVA tests were used to determine the attributes of CPGs associated with the total score in AGREE II. Ten CPGs published between 2010 and 2020 were included, mostly from Europe (n=6). The overall agreement between reviewers was very good (0.94; 95%CI 0.91-0.97). The mean scores for each domain were as follows: Scope and purpose 78.0 ± 18.9%; Stakeholder involvement 46.9 ± 29.6%; Rigour of development 41.8 ± 26.7%; Clarity of presentation 75.8 ± 17.6%; Applicability 15.3 ± 18.8% and Editorial independence 41.7 ± 41.7%. The overall mean rate was 4 ± 1.3 out of a maximum score of 7. Only two guidelines were recommended by the reviewers and rated as high quality. The CPGs developed by governments showed a significantly higher overall score. The overall quality of CPGs on TDI was suboptimal. Therefore, the CPGs developers need to use a methodology that allows them to formulate recommendations in a structured, transparent, and explicit way.

## INTRODUCTION

A traumatic dental injury (TDI) is an impact injury that affects the tooth and the supporting structures surrounding it [1]. TDIs are a serious public health problem due to their prevalence and their consequences for the quality of life of the affected patients [2]. The estimated prevalence of TDIs worldwide is 22.7% in primary teeth and 15.2% in permanent teeth, with an estimated global incidence rate of 2.82 (number of events per 100 persons per year) [3]. The study by Petti *et al*. (2018) on the global burden of TDIs shows that more than one billion people have had at least one TDI; and if ranked as an acute/chronic disease and injury, it would be positioned as the 5^th^ most prevalent condition worldwide [3].

Proper diagnosis of TDIs, together with treatment planning and follow-up, are fundamental for ensuring a favorable outcome and prognosis for the affected teeth and patients [4]. Nevertheless, this task is not easy to achieve because of the complexity of diagnosing TDIs and the multiple treatment options available. A recent systematic review showed insufficient knowledge of TDI prevention and emergency management by dental professionals worldwide [5]. This lack of expertise induces a significant variability in the management of TDIs, impacting directly the patient’s oral health and quality of life [6], along with high costs for health systems [7; 8].

One way to help clinicians to make a proper diagnosis, guide them to the most appropriate therapy and reduce clinical variability is through clinical practice guidelines (CPGs). CPGs are developed by clinical experts, drawing up evidence-based recommendations to help health professionals and patients to make an appropriate decision in specific clinical circumstances [9].

Evidence shows that CPGs of different dentistry specialties tend to be assessed as low quality, specially related to lack of methodological rigor in their development [10; 11] and problems in applicability [12; 13], making their implementation unreliable and their use difficult for patients, clinicians, and policy-makers. Poor quality CPGs may influence negatively the patient care or have a debatable applicability [14; 15].

So far as we can tell, there is no systematic quality assessment of CPGs for TDIs; therefore, little is known about their quality, potential impact, and applicability. The aim of this study was to identify and evaluate the quality of CPGs for the diagnosis, emergency management and follow-up of TDIs.

## MATERIALS AND METHODS

### Study Design

We carried out a systematic quality evaluation of CPGs on TDIs using the AGREE II tool and following a methodology published previously [10; 13]. The protocol was pub-lished in the Open Science Framework [16].

### Eligibility Criteria

Documents published in English, German, Portuguese, and Spanish were included, that were self-declared as a guideline, or provided recommendations for the emergency management or treatment of the consequences of TDIs. We only included the most recent version of the identified CPGs. Documents without recommendations and discontinued CPGs were excluded.

### Sources of Information

We made a systematic search in the MEDLINE (via PubMed), EMBASE, Episte-monikos and Trip (Turning Research Into Practice) databases up to May 22, 2021. Guideline developers’ websites, repositories, Health Ministries and international dental scientific societies were also screened. This search was also updated in May of 2022. We did not limit the search by date, language, or publication status. The details of the search strategy can be found in the supplementary data (Appendix S1).

### Selection of the Guidelines

The titles, abstracts, and full texts were reviewed independently by 2 researchers (R.D., A.S.) in a 3-step process using Rayyan® software (www.rayyan.ai). If there was a discrepancy, a third reviewer resolved it (C.Z.).

### Data Charting Process

Two reviewers (A.S., C.A., G.E. or N.F.D.) extracted the following characteristics of each CPG independently: author, year, title, country, organization, language, scope (emergency or treatment), target population, method used for the quality assessment and recommendation development methodology of the studies included.

### Critical Appraisal of Individual Sources of Evidence

Two reviewers (C.A., C.Z., G.E., or N.F.D.) worked independently to rate the quality of each guideline with the AGREE II instrument (https://www.agreetrust.org/resource-centre/agree-plus/). AGREE II consists of 23 items and six domains: 1. Scope and purpose; 2. Stakeholder involvement; 3.Rigour of de-velopment; 4. Clarity of presentation; 5. Applicability; and 6. Editorial independence. Each item is rated on a Likert scale ranging from 1 (lowest quality) to 7 (highest quality) points. AGREE II includes two overall quality ratings for each CPG: i) an overall score of 1 to 7 and ii) a reviewer recommendation classing it as “recommended”, “recommended with modifications”, or “not recommended” [17].

### Data Analysis

The total AGREE II score was calculated by totaling the scores of all the individual items in each domain, and then scaling the total score as a percentage of the highest possible score for the domain [17]. Discrepancies between reviewers that exceeded 3 points, or standard deviation (SD) in any item equal to or greater than 1.5 SD, were reassessed [10; 13]. The standardized score was calculated for each domain (range 0 to 100%) [17]. CPGs with a score of 60% or higher in at least 3 domains, including Rigour of de-velopment, were classified as high-quality [10; 11; 13].

Overall agreement among the reviewers was calculated using the intraclass coeffi-cient with 95% confidence interval (95% CI). Agreement between 0.01 and 0.20 was considered slight, from 0.21 to 0.40 fair, from 0.41 to 0.60 moderate, from 0.61 to 0.80 substantial, and from 0.81 to 1.00 very good [18].

T-student or ANOVA tests were used to determine associations between the total score in AGREE II and the attributes of the CPGs, e.g. year of development (last five years or more), CPGs development agencies (Government, Scientific societies, or hospitals), and region (Europe, America, Asia). Any significant ANOVA was checked by post-hoc tests (Tukey’s Honestly Significant Differences) to determine differences between groups.

Finally, we used the Pearson correlation coefficient to evaluate correlations between the AGREE II domain scores and the total score, in order to determine which domains influenced the overall quality of the CPG. Pearson’s correlation was interpreted as follows: r <0.1 negligible, 0.1 - 0.39 weak, 0.4 -0.69 moderate, 0.7-0.89 strong, and r >0.8 very strong [19].

## RESULTS

The selection flow chart is shown in Figure 1. The systematic search retrieved 479 articles, and other sources identified 80 documents/articles. After excluding duplicates and studies that failed to meet the inclusion criteria, ten CPGs were included in the final analysis.

**Figure 1.**
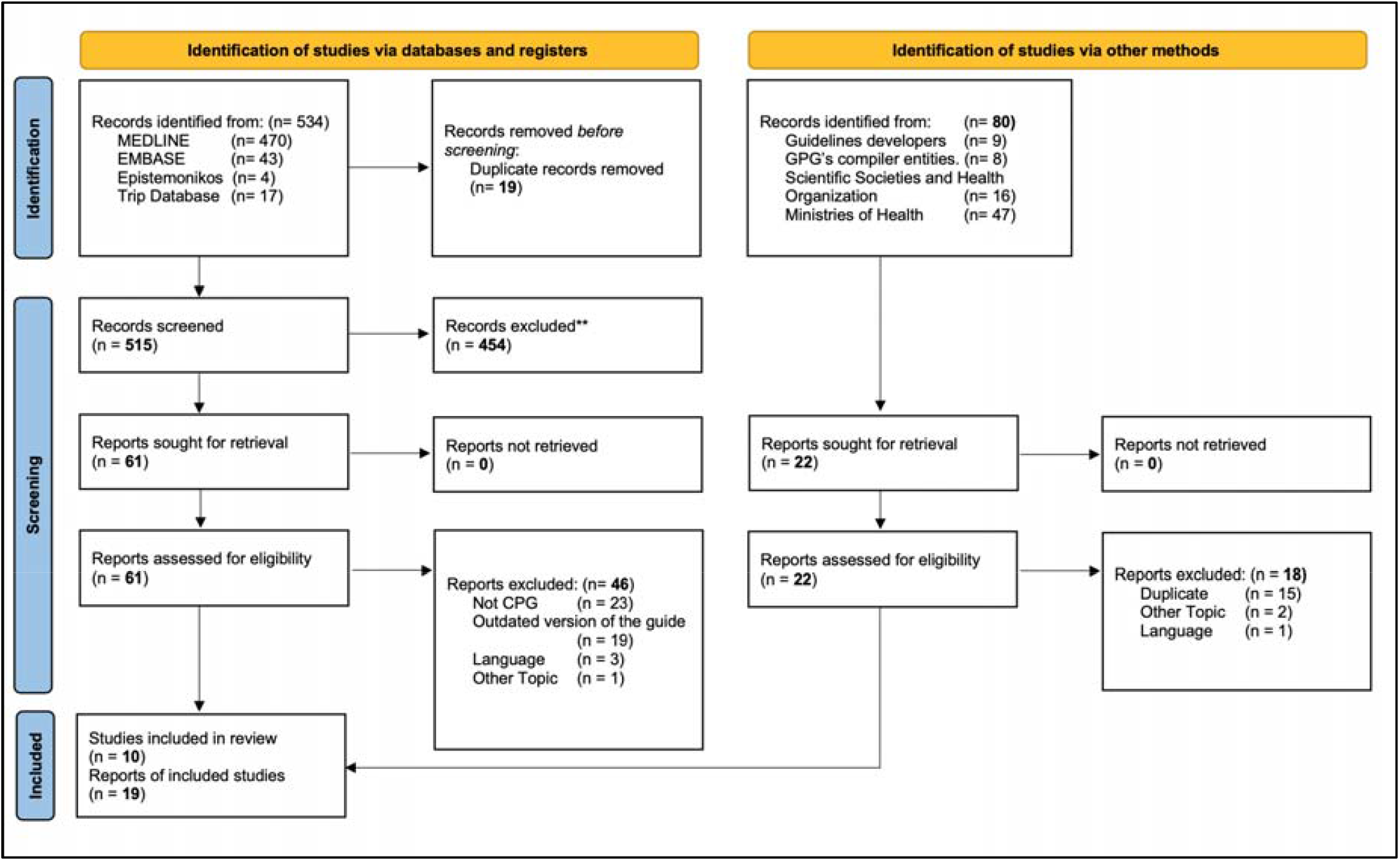
Flow-chart of the selection process.

Table 1 lists the characteristics of the CPGs, which were all published between 2010 and 2022. Eight were in English: three from UK and one each from Italy, Norway, Malaysia and USA, and one global. Of the other two, one was in Spanish and the other in German. Six CPGs were focused on recommendations for the management of all types of TDIs, two for avulsed teeth, one for intruded teeth and one for endodontic management of traumatized permanent teeth. The developers were scientific societies or dental colleges (n=6), Ministries or government agencies (n=3) and one from a Hospital. Only two CPGs reported their funding source. Only three of the CPGs [4; 20-25] are recent updates from a previous version. Although most of the guidelines declared that systematic methods were used for the search of evidence (n=9), only two assessed the risk of bias [26; 27]. Most of the guidelines that reported a methodology for the drafting of the recommendations did so through consensus [26].

**Table 1.**
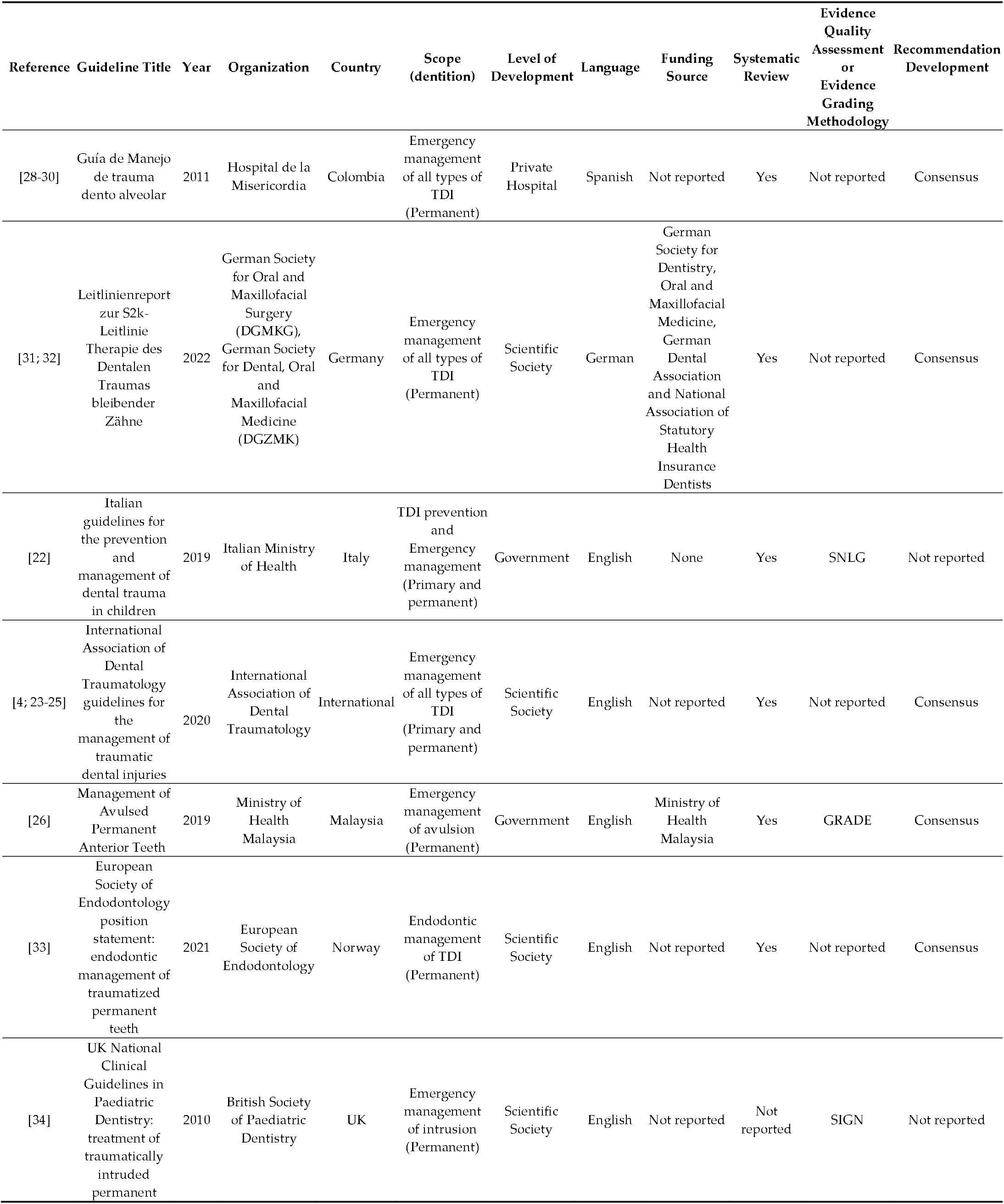
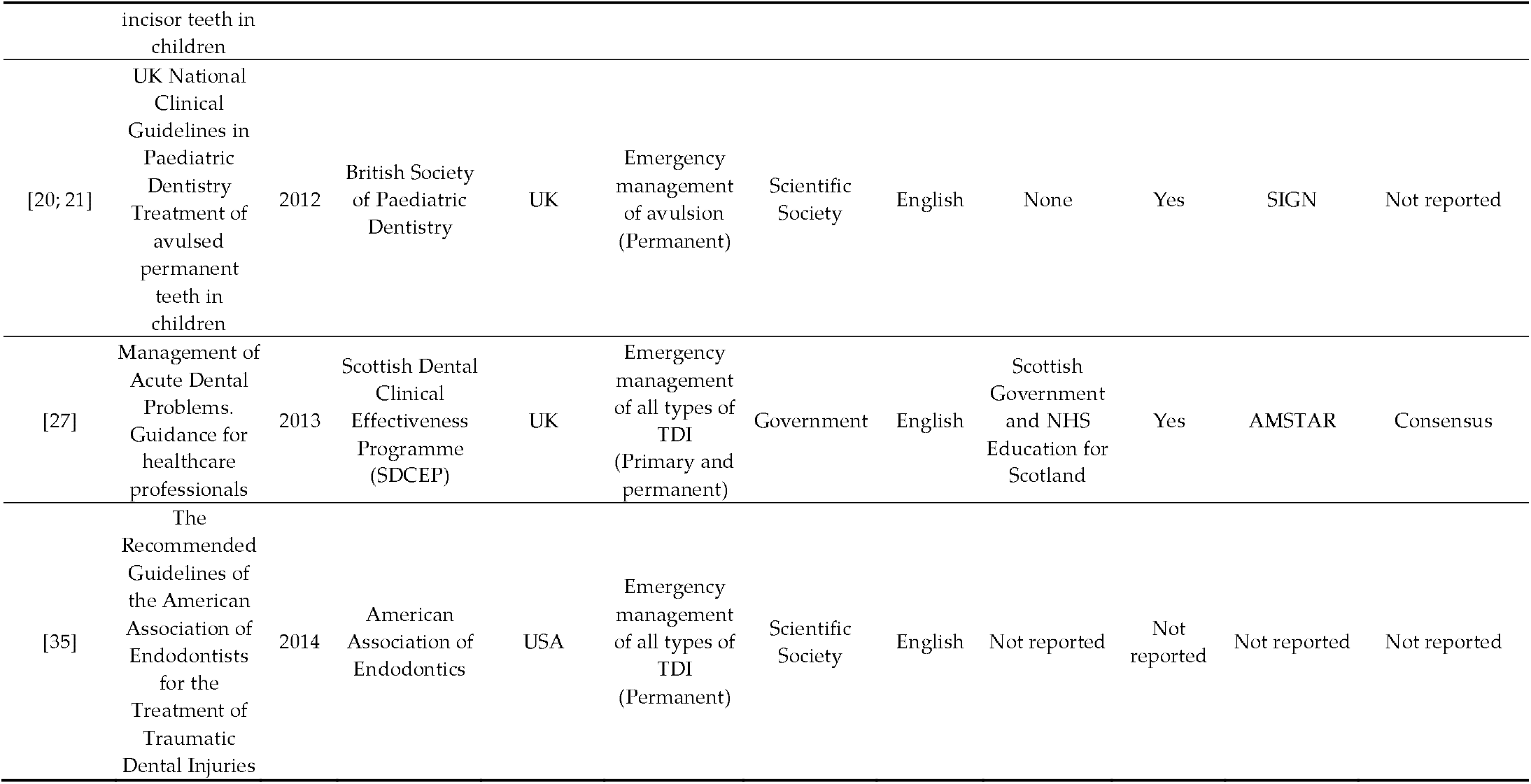
Characteristics of the guidelines included.

### Appraisal of CPGs

Overall, agreement between the reviewers was classed as very good (ICC= 0.94; 95%CI 0.91-0.97). **Table 2** shows the standardized scores for each CPG by domain, and the overall recommendation. The only domains to score above 60% were Scope and purpose and Clarity of presentation. The domain with the lowest score was Applicability, with a mean of 15.3% ± 18.8.

**Table 2.** Standardized scores of guidelines by domain

| Reference | Guideline | Scope and purpose | Stakeholder involvement | Rigor of development | Clarity of presentation | Applicability | Editorial independence | Overall rate | Overall Recommendation | Quality |
| --- | --- | --- | --- | --- | --- | --- | --- | --- | --- | --- |
| [28-30] | Guía de Manejo de trauma dento alveolar | 50% | 22% | 15% | 36% | 2% | 0% | 2 | Not recommended | Low |
| [31; 32] | Leitlinienreport zur S2k-Leitlinie Therapie des Dentalen Traumas bleibender Zähne | 97% | 83% | 51% | 78% | 10% | 100% | 4.5 | Recommended with modifications | Low |
| [22] | Italian guidelines for the prevention and management of dental trauma in children | 100% | 67% | 82% | 83% | 0% | 96% | 6 | Recommended | High |
| [4; 23-25] | International Association of Dental Traumatology guidelines for the management of traumatic dental injuries | 69% | 17% | 34% | 86% | 15% | 25% | 4.5 | Recommended with modifications | Low |
| [26] | Management of Avulsed Permanent Anterior Teeth | 92% | 75% | 52% | 64% | 56% | 88% | 5 | Recommended with modifications | Low |
| [33] | European Society of Endodontology position statement: endodontic management of traumatized permanent teeth | 75% | 33% | 33% | 92% | 0% | 50% | 3.5 | Not recommended | Low |
| [34] | UK National Clinical Guidelines in Paediatric Dentistry: treatment of traumatically intruded permanent incisor teeth in children | 58% | 33% | 20% | 69% | 13% | 4% | 2.5 | Not recommended | Low |
| [20; 21] | UK National Guidelines in Paediatric Dentistry Treatment of avulsed permanent teeth in children | 100% | 50% | 55% | 94% | 17% | 0% | 4.5 | Recommended with modifications | Low |
| [27] | Management of Acute Dental Problems. Guidance for healthcare professionals | 81% | 86% | 74% | 67% | 40% | 54% | 5 | Recommended | High |
| [35] | The Recommended Guidelines of the American Association of Endodontists for the Treatment of Traumatic Dental Injuries | 58% | 3% | 2% | 89% | 0% | 0% | 2.5 | Not recommended | Low |
| Mean |  | 78.0% | 46.9% | 41.8% | 75.8% | 15.3% | 41.7% | 4.0 |  |  |
| (SD) |  | 18.9% | 29.6% | 25.7% | 17.6% | 18.8% | 41.7.% | 1.3 |  |  |
| (Minimum–Maximum) |  | (50-100%) | (3-86%) | (2-82%) | (36-94%) | (0-56%) | (0-100%) | 2 - 6 |  |  |

### Scope and Purpose

The mean score was 78.0% ± 18.9 (range 50-100%). Of the 10 guidelines, seven scored above 60% in this domain, demonstrating that most of the guidelines defined well the target audience for whom the CPG was planned.

### Stakeholder Involvement

Four CPGs scored above 60% in the Stakeholder involvement domain, while the mean score was 46.9% ± 29.6 (range 3-86%). The main limitations of this domain were the need for more detailed information about the group that developed the guideline (discipline, institution, description of role) and failure to consider the preferences of target users’.

### Rigour of Development

For this domain, the mean score was 41.8% ± 25.7 (range 2-82%). Only two guidelines scored above 60%. Although most of the guidelines declared that they had conducted a systematic search of evidence, only two formally assessed the strengths and limitations of the supporting evidence [26; 27]. However, three guidelines graded the evidence of the included studies in an effort to assess the quality of the supporting evidence [20; 21; 34] [22].

Scarce information was provided on the methods used to develop the recommendations. However, most CPGs used consensus as the method for the panel members to reach their decisions. Seven guidelines reported a direct link between the supporting evidence and the recommendations. Four CPGs reported information on external peer review prior to dissemination [20-22; 26; 27] and 2 reported appropriate information about the updating process [26; 27].

### Clarity of Presentation

In this domain, the mean score was 75.8% ± 17.6% (range 36-94%). Only one CPG scored below 60% in this domain, indicating that the recommendations were clearly presented.

### Applicability

All the guidelines scored less than 60% in the Applicability domain. The mean score for this domain was 15.3% ± 18.8 (range 0-56%). The main limitations were that most of the CPGs did not discuss barriers to and facilitators of implementation, did not evaluate the implications in the use of resources, or did not present key review criteria for the purposes of monitoring and/or auditing [26; 27].

### Editorial Independence

For this domain, the mean score was 41.7% ± 41.7 (range 0-100%). Seven CPGs scored below 60% and two of them scored 0.0%. Some CPGs did not describe fully their sources of funding and the possible influence of these on CPG development, or failed to report the authors’ potential conflicts of interest.

### Overall Assessment

Only two of the guidelines were classed by the reviewers as recommended, and four were recommended with modifications. After the assessment, two of the CPGs were classed as high quality (scored ≥ 60% in at least three domains, including Rigour of Development). The overall mean was 4 ± 1.3, the highest score awarded was 6, while the lowest was 2.

### Association Between AGREE II Score and Characteristics of the CPGs

The CPGs developed by governments showed a significantly higher overall score than the guidelines published by scientific societies or hospitals. Nonetheless, this difference was not substantial across any domain except for the Clarity of presentation. We found no significant differences between the guidelines that were developed in the last five years or earlier, and between the continents where the CPG was developed. However, the CPGs developed in Asia were better at reporting the aspects related to Applicability, and the most recent CPGs stated Editorial independence more clearly (**Table 3**).

**Table 3.** Comparison between AGREE II domains and pre-specified predictors.

| Variables | *n* | Scope and purpose | Stakeholder involvement | Rigour of development | Clarity of presentation | Applicability | Editorial Independence | Overall rate |
| --- | --- | --- | --- | --- | --- | --- | --- | --- |
|  |  | Mean (SD) | Mean (SD) | Mean (SD) | Mean (SD) | Mean (SD) | Mean (SD) | Mean (SD) |
| **Year of development** |  |  |  |  |  |  |  |  |
| ≥ 2018 | 5 | 86.6 (13.8) | 55.0 (28.5) | 50.4 (19.8) | 80.6 (10.6) | 16.2 (23.2) | 71.8 (32.8) | 4.7 (0.9) |
| < 2018 | 5 | 69.4 (9.2) | 38.8 (31.4) | 33.2 (30.1) | 71.0 (22.9) | 14.4 (16.0) | 11.6 (23.8) | 3.3 (1.4) |
| p-value |  | 0.106 | 0.418 | 0.317 | 0.420 | 0.890 | 0.011* | 0.091 |
| **Type of organization** |  |  |  |  |  |  |  |  |
| Government | 3 | 91.0 (9.5) | 76.0 (9.5) | 69.3 (15.5) | 71.3 (10.2) | 32.0 (28.8) | 79.3 (22.3) | 5.33 (0.77) |
| Scientific societies | 6 | 76.2 (18.5) | 36.5 (27.8) | 32.5 (19.7) | 84.7 (9.5) | 9.2 (7.5) | 29.8 (39.5) | 3.67 (0.98) |
| Hospital | 1 | 50.0 (0.0) | 22.0 (0.0) | 15.0 (0.0) | 36.0 (0.0) | 2.0 (0.0) | 0.0 (0.0) | 2.0 (0.0) |
| p-value |  | 0.159 | 0.098 | 0.043* | 0.007* | 0.178 | 0.134 | 0.027* |
| **Region** |  |  |  |  |  |  |  |  |
| Asia | 1 | 92.0 (0.0) | 75.0 (0.0) | 52.0 (0.0) | 64.0 (0.0) | 56.0 (0.0) | 88.0 (0.0) | 4.35 (1.11) |
| America | 2 | 54.0 (5.7) | 12.5 (13.4) | 8.5 (9.2) | 62.5 (37.5) | 1.0 (1.4) | 0.0 (0.0) | 2.25 (0.35) |
| Europa | 7 | 82.6 (16.6) | 52.7 (26.7) | 49.9 (22.6) | 81.3 (10.5) | 13.6 (13.5) | 47.0 (40.4) | 5.0 (0.0) |
| p-value |  | 0.107 | 0.139 | 0.110 | 0.359 | 0.024* | 0.196 | 0.08 |

\* Statistically significant difference.

A strong significant correlation was observed between the scores of the AGREE II domains and the overall rate, excepting the Clarity of presentation (*r=*0.32; p=0.363) and Applicability domains (*r*=0.43; p=0.21) (**Table 4**).

**Table 4.** Correlation score between the scores for each AGREE II instrument domain and the overall.

| Analysis | Scope and purpose | Stakeholder involvement | Rigour of development | Clarity of presentation | Applicability | Editorial Independence |
| --- | --- | --- | --- | --- | --- | --- |
| *r* | 0.8722 | 0.7319 | 0.925 | 0.342 | 0.421 | 0.743 |
| p-value | <0.001* | 0.016* | <0.001* | 0.333 | 0.226 | 0.014* |

\* Statistically significant difference.

## DISCUSSION

### Main Findings

Our research showed that the overall quality of CPGs in the field of dental trauma is suboptimal; only two out of ten CPGs were assessed as high quality. The domain with the highest score was Scope and Purpose (mean 78.0%), while Applicability obtained the lowest score (mean 15.3%). The AGREE II overall mean rate was 4.0 (SD 1.3). Only two CPGs were recommended without modifications by the reviewers.

The only studied variable associated with the quality of the guidelines was the organization in charge of developing the guidelines, since the CPGs developed by governments were found to present the best quality. Finally, as expected, the best correlated domain with a high-quality CPG was Rigour of development.

### Results of this work in the Context of Previous Research

Our review showed that the two best assessed domains are Scope and purpose and Clarity of presentation, consistent with other systematic reviews [36; 37].

Although the Scope and purpose domain passed the quality threshold, some guidelines failed to describe the health questions covered by the CPG. Well formulated study questions help directly the search for evidence, as well as the assessment of certainty; therefore, when choosing which questions to include, the objective and scope of the guide are being defined [38]. Since the recommendations are the answer to these questions, the object of the CPG should be clear, and consistent with the recommendations, in order to help the user to implement the most appropriate care for a given patient.

The Clarity of presentation domain presented the second-best evaluation, the main issues being ambiguity and the format in which the recommendations were presented. This is relevant for making the recommendation easier to implement.[17].

As in our study, evidence shows that dental CPGs of different dental specialties tend to be of low quality, presenting important flaws in their development, especially related to Stakeholder involvement, poor Methodological rigor [10; 39], and issues in the Applicability and the Editorial independence domains [12; 13].

Regarding the Stakeholder involvement domain, the views and preferences of the target population were not considered in formulating the recommendations, either because they were not included as members of the panel or because the study did not carry out a systematic search of the evidence. The principal justification for including patients’ values and preferences in guideline development is because recommendations that are in line with these might be more easily accepted, implemented and adhered to by those who will benefit from them. [40]. Moreover, most CPGs should have stated the specialists or experts involved in their development. CPGs improve when specialists, methodologists and patients participate actively in guideline development [41; 42].

Rigour of development is regarded as the most important domain for the assessment of CPGs, since it appraises the process for the gathering and synthesizing the evidence, and the methodology for formulating the recommendations. Although most of the CPGs reported that they carried out a systematic search of the evidence, few of them assessed the strengths and limitations of the evidence collected. This is important since most of the evidence that supports the recommendations comes from observational or animal studies. The certainty of the evidence in these cases is low or very low, determining a conditional or weak recommendation, meaning that some individuals in this position might accept the suggested course of action, but that many would not [43].

Another important limitation in the methodological rigor of the CPGs was that the methodology for formulating the recommendations was not clearly described. Although most of the CPGs reported that the recommendations were formulated by consensus of the panel members, few provided information on the methodology, the factors considered and the results of the deliberation process. One way to make this process more transparent is through the GRADE approach (Grading of Recommendation Assessment, Development and Evaluation). This methodology provides a structured process for determining the certainty of the evidence, making recommendations, and taking decisions. The GRADE approach does not only consider the quality of the evidence when formulating a recommendation, but also considers the benefit-risk balance, the patients’ values and preferences, the magnitude of the necessary resources and costs, as well as equity, acceptability, and implementation, among others. Evidence shows that the best quality CPGs are those based on evidence and their use is a transparent way to develop recommendations, backed by the GRADE methodology [12]. This is important given that poor quality guidelines may negatively influence patient care, or their applicability may be questionable [14; 15]. In our study, only one guideline used the GRADE approach to assess the certainty of the evidence and to develop its recommendations, despite the fact that more than 90 health organizations around the world have endorsed this approach [44]. However, this is also a deficiency seen in CPGs of other dentistry areas [11; 13; 45].

The Applicability domain is poorly reported in the CPGs, not only in dental guidelines, but also in other health fields [37]. This shows the importance of considering aspects such as implementation, organizational barriers and facilitators, and economic implications when developing future guidelines on TDIs. Inappropriate analysis of these factors can influence adherence to the guideline. When carrying out this analysis, the CPGs must consider the local facilitators and the barriers that may influence their applicability. According to Alonso-Coello et al., low scores in the applicability domain could result from the fact that the developers consider guideline development and guideline implementation as different activities [37].

The Editorial Independence domain was assessed as very low-quality because the CPGs did not declare possible intellectual and financial conflicts of interest. This is a cross-cutting problem, both in dental and medical guidelines [36; 37] [46]. It is essential that both funding bodies and members of CPG development groups state their conflicts in detail, because CPGs are used for decision-making in both insurance coverage and standards of care [47]. It is important to link the recommendations clearly to the evidence, and to exclude panelists with conflicts of interest, in order to avoid influence from external interests [46].

Concerning the factors associated with guideline quality, we observed that guidelines developed by Governments have higher scores than CPGs produced by scientific societies or hospitals, in agreement with the information reported in other studies [37; 48]. This is based on the large amount of financial and human resources needed to properly develop a CPG [49].

### Strengths and Limitations

The greatest strength of the present study was that CPGs were obtained by a systematic literature search that included developers’ websites and repositories of CPGs. AGREE II is the only reliable, validated instrument developed for comparing CPGs. [17].

Our study is not exempt from limitations. Although a comprehensive search including gray literature was conducted, relevant guidelines may exist in a language other than those considered in our methodology. Likewise, it is important to note that the recommendations of the CPGs assessed should be viewed with caution, since AGREE II only assesses the reporting of aspects of methodological quality, without judging the rationality of the recommendations made. Other approaches, such as GRADE, should be used to assess the certainty of evidence supporting the recommendations.

### Implications for Practice and Investigation

One of the principal implications of our study is that it evidences the need to improve the development processes for CPGs in dental traumatology, since dentists should identify trustworthy CPGs before implementing the recommendations. The development groups should focus on using basic quality criteria for CPG development, such as description of the methodology used, assessing the quality of the body of evidence and recommendations, as well as the process by which consensus is reached by the members of the panel of experts. Quality improvements should be pursued by using a transparent and standardized framework for presenting the recommendations, and by considering a balance between the desirable and undesirable effects of the interventions, the patients’ values and preferences in the development process, the certainty of evidence for the resources required, the cost-effectiveness of the interventions, and other contextual aspects such as the impact on health equity and the acceptability or feasibility of implementation.

Since developing trustworthy guidelines requires substantial investment in time and resources, adaptation or adoption of existing high-quality guidelines may be considered as an alternative to developing new guidelines, increasing the efficiency of guideline development.

## Data Availability

All data produced in the present study are available upon reasonable request to the authors

## CONCLUSION

The overall quality of CPGs for the diagnosis, emergency management, and follow-up of TDIs was suboptimal, with only two high quality guidelines out of the ten assessed, making implementation challenging for dentists and policy makers. It is essential that guideline developers should use a methodology that allows them to formulate the recommendations in a structured, transparent, and explicit way.

## Supplementary Materials

Supplementary Material S1. Search strategies used in the databases.

## Author Contributions

Conceptualization, A.S., C.A., C.Z., and R.D..; methodology, A.C.L. and C-Z..; validation, C.Z., and G.E.E.; formal analysis, A.S., C.A., C.Z., G.E.E., and R.D.; investigation, A.S., C.A., G.E.E.; data curation, C.A., C.Z., and N.F.D.; writing—original draft preparation, C.Z., A.C.L.; writing—review and editing, and A.C.L.; C.Z., and N.F.D., visualization, A.C.L., and C.Z..; supervision, C.Z..; project administration, C.Z.

## Funding

This research received no external funding.

## Institutional Review Board Statement

Not applicable.

## Informed Consent Statement

Not applicable.

## Data Availability Statement

Not applicable.

### Acknowledgments

Not applicable.

## Conflicts of Interest

The authors declare no conflict of interest.

## Notes

### Competing Interest Statement

The authors have declared no competing interest.

