## Supplementary material for "Emergency and Sequalae Management of Traumatic Dental Injuries: A Quality Assessment of Clinical Practice Guidelines"

**APPENDIX S1**

**SEARCH STRATEGY USED IN EACH DATABASE**

| **Source** | **Strategy** | **Results** |
| --- | --- | --- |
| Medline | (((((((((tooth[tiab]) OR teeth) OR dental[tiab]) OR dentoalveolar)) AND (((((injur*[tiab]) OR traumatology[tiab]) OR "Traumatology"[Mesh]) OR ("Wounds and Injuries"[Mesh])) OR trauma[tiab]))))) AND ((((((((((((guideline*[ti]) OR recommendation*[ti]) OR protocol*[ti]) OR consensus[ti]) OR practice guideline[ti]) OR guidance[ti]) OR "Guideline" [Publication Type]) OR "Practice Guideline" [Publication Type]) OR "Health Planning Guidelines"[Mesh]) OR "Consensus"[Mesh]) OR  "Guidelines as Topic"[Mesh])) | 470 |
| Embase | (guideline*:ti OR 'guideline'/exp OR 'practice guideline'/exp OR 'consensus development'/exp OR 'consensus'/exp OR recommendation*:ti OR consensus:ti OR protocol*:ti OR 'protocol'/exp) AND ('traumatic dental injur*':ti,ab OR 'dental trauma':ti,ab OR 'dentoalveolar trauma':ti,ab  OR 'tooth injur*':ti,ab OR 'teeth injur*':ti,ab) AND [embase]/lim | 43 |
| Epistemonikos | (title:(traumatic dental injur*) OR abstract:(traumatic dental injur*)) OR (title:(dental trauma) OR abstract:(dental trauma)) OR (title:(dentoalveolar trauma) OR abstract:(dentoalveolar trauma)) OR (title:(tooth injur*) OR abstract:(tooth injur*)) OR (title:(teeth injur*) OR  abstract:(teeth injur*)) [Filters: protocol=no, classification=broad- synthesis] | 4 |
| TripDatabase | “traumatic dental injur*” OR “dental trauma” OR “dentoalveolar trauma” OR “tooth injur*” OR “teeth injur*” | 17 |

**WEBSITES**

### Guidelines developers

- National Institute for Health and Care Excellence (http://www.nice.org.uk)
- Scottish Intercollegiate Guidelines Network (http://www.sign.ac.uk)
- Guía Salud (https://portal.guiasalud.es)
- CMA Infobase: Clinical Practice Guidelines Database (CPGs) (https://joulecma.ca/cpg/homepage)
- Australian Clinical Practice Guideline (https://www.clinicalguidelines.gov.au)
- New Zealand Guidelines (https[://www.](http://www.health.govt.nz/about-ministry/ministry-)hea[lth.govt.nz/about-ministry/ministry-](http://www.health.govt.nz/about-ministry/ministry-) health-websites/new-zealand-guidelines-group)
- Scottish Dental Clinical Effectiveness Programme (http://www.sdcep.org.uk)
- EBM Guidelines (https[://www](http://www.ebm-guidelines.com/dtk/ebmg/home)).eb[m-guidelines.com/dtk/ebmg/home)](http://www.ebm-guidelines.com/dtk/ebmg/home))
- IECS: Instituto de Efectividad Clínica y Sanitaria (https://www.iecs.org.ar).

### CPGs compiler entities

- National Guideline Clearinghouse (http://www.guideline.gov)
- Guideline International Networks (GIN): (<http://www.g-i-n.net/library/international-guidelines-> library)
- ECRI Guidelines Trust (https://guidelines.ecri.org/)
- NeLH Guidelines Finder (<http://libraries.nelh.nhs.uk/guidelinesFinder/)>
- Agency for Healthcare Research and Quality (AHRQ) https[://www.](http://www.ahrq.gov/research/findings/evidence-based-reports/search.html))ahr[q.gov/research/findings/evidence-based-reports/search.html)](http://www.ahrq.gov/research/findings/evidence-based-reports/search.html))
- Guideline Central(https[://www.](http://www.guidelinecentral.com/))gu[idelinecentral.](http://www.guidelinecentral.com/))c[om/)](http://www.guidelinecentral.com/))
- The Alliance for the Implementation of Clinical Practice Guidelines (https://aicpg.org/)
- Scottish dental (https[://www.](http://www.scottishdental.org/professionals/guidelines/))sco[ttishdental.org/professionals/guidelines/)](http://www.scottishdental.org/professionals/guidelines/))

### Scientific Societies and Health Organizations

- World health Organization (OMS) (https[://www.](http://www.who.int/es))who[.int/es)](http://www.who.int/es))
- Organización Panamericana de la Salud (OPS) (https[://www](http://www.paho.org/en)).p[ah](http://www.paho.org/en))o[.org/en)](http://www.paho.org/en))
- American Dental Association (ADA) (https[://www.](http://www.ada.org/en))ad[a.](http://www.ada.org/en))o[rg/en)](http://www.ada.org/en))
- FDI World Dental Association (https://www.fdiworlddental.org)
- International Association for Dental Research (IADR)(https://www.iadr.org)
- American Academy of Pediatric Dentistry (AAPD) (<http://www.aapd.org/)>
- Australian and New Zealand Society of Pediatric Dentistry (ANZSPD) (https[://www.](http://www.anzspd.org.au/))an[zsp](http://www.anzspd.org.au/))d[.org.au/)](http://www.anzspd.org.au/))
- European Academy of Paediatric Dentistry (EAPD) (<http://www.eapd.gr/)>
- International Association of Paediatric Dentistry (IAPD) (<http://www.iapdworld.org/)>
- Pediatric Dentistry Association of Asia (PDAA) (<http://pdaasia.org/>)
- International association of dental traumatology ([https://www.iadt-dentaltrauma.org](https://www.iadt-dentaltrauma.org/))
- International association of oral and maxillofacial surgeons ([https://www.iaoms.org](https://www.iaoms.org/))
- International Association of DentoMaxilloFacial Radiology (IADMFR) ([https://iadmfr.one](https://iadmfr.one/))
- International Federation of Endodontic Associations (<http://www.ifeaendo.org/about-us/what-is-> ifea/)
- American association of endodontists ([https://www.aae.org](https://www.aae.org/))
- European society of endodontology ([https://www.e-s-e.eu](https://www.e-s-e.eu/))

### Ministries of Health

- Africa: Nigeria (http://www.fmh.gov.ng), South Africa ([http://www.doh.gov.za/),](http://www.doh.gov.za/)) Sudan ([http://www.fmoh.gov.sd/),](http://www.fmoh.gov.sd/)) Tanzania (<http://www.moh.go.tz/)> and Uganda ([http://health.go.ug/mohweb/).](http://health.go.ug/mohweb/))
- America: Argentina (https[://www.](http://www.argentina.gob.ar/salud))arg[entina.gob.ar/salud),](http://www.argentina.gob.ar/salud)) Brasil (https://saude.gov.br), Bolivia (https://www.minsalud.gob.bo), Chile (http://www.minsal.cl), Colombia (https[://www.](http://www.minsalud.gov.co/portada-covid-)min[salud.gov.co/portada-covid-](http://www.minsalud.gov.co/portada-covid-) 19.html), Costa Rica ([http://www.ministeriodesalud.go.cr/),](http://www.ministeriodesalud.go.cr/)) Cuba ([http://www.sld.cu/),](http://www.sld.cu/)) Dominican Republic ([http://www.salud.gob.do/),](http://www.salud.gob.do/)) El Salvador ([http://www.salud.gob.sv/),](http://www.salud.gob.sv/)) Nicaragua and ([http://www.minsa.gob.ni/),](http://www.minsa.gob.ni/)) Perú (https[://www.](http://www.gob.pe/minsa/))gob[.pe/minsa/),](http://www.gob.pe/minsa/)) Trinidad and Tobago (<http://www.health.gov.tt/)> and United States of America ([http://www.hhs.gov/).](http://www.hhs.gov/))
- Asia: Bhutan ([http://www.health.gov.bt/),](http://www.health.gov.bt/)) Cambodia ([http://www.moh.gov.kh/),](http://www.moh.gov.kh/)) China (http://www.moh.gov.cn), Indonesia (http://www.depkes.go.id), Iraq (http://www.moh.gov.iq), Japon (<http://www.mhlw.go.jp/>), Pakistan (www.pakistan.gov.pk), Singapore

([http://www.health.gov.lk/),](http://www.health.gov.lk/)) Thailand (<http://eng.moph.go.th/)> and Turkey ([http://www.sb.gov.tr/).](http://www.sb.gov.tr/))

- Europe: Austria ([http://www.bmg.gv.at/),](http://www.bmg.gv.at/)) Belgium (http://www.health.belgium.be), Cyprus (http://www.moh.gov.cy), Denmark ([http://www.sst.dk/),](http://www.sst.dk/)) England (http://www.dh.gov.uk), France ([http://www.sante.gouv.fr/),](http://www.sante.gouv.fr/)) Germany (http://www.bmg.bund.de), Greece ([http://www.yyka.gov.gr/),](http://www.yyka.gov.gr/)) Iceland ([http://www.velferdarraduneyti.is/),](http://www.velferdarraduneyti.is/)) Ireland (http://www.dohc.ie), Italy ([http://www.salute.gov.it/),](http://www.salute.gov.it/)) Netherlands ([http://www.government.nl/ministries/vws),](http://www.government.nl/ministries/vws)) Norway (http://www.regjeringen.no), Portugal (http://www.portaldasaude.pt), Spain ([http://www.msc.es/),](http://www.msc.es/)) Sweden (<http://www.folktandvardenstockholm.se/)> and Switzerland (http://www.bag.admin.ch).
- Oceania: Australia (<http://www.health.gov.au/)> and New Zealand (http://www.health.govt.nz).
